# Functional Effect Predictions For Ion Channel Missense Variants Using a Protein Language Model

**DOI:** 10.1101/2025.10.16.25337735

**Authors:** Sean Gies, Artoghrul Alishbayli, Paul H.E. Tiesinga, Marijn B. Martens

**Affiliations:** Synaptica Ltd, Nijmegen, The Netherlands; Dept of Neurophysics, Radboud University, Nijmegen, The Netherlands

**Author notes:** These authors contributed equally to this work (Sean Gies, Artoghrul Alishbayli).

**Keywords:** channelopathy, gain-of-function, loss-of-function, variant functional effect, precision medicine, rare diseases

## Abstract

Channelopathies represent a group of diseases often caused by missense variants in ion channels affecting the functioning of tissues like the nervous system, heart, and muscle. The gold standard for functionally characterizing a variant is to measure the electrophysiological changes in channel properties using cell-based heterologous expression systems. As this method is time-consuming and generally unavailable, clinical practice often relies on *in silico* models to predict the functional consequences of ion channel variants. We constructed a Missense ION (MissION) channel variant classifier based on a protein language model and trained on 3176 gain- or loss-of-function variants, the largest set collected to date, in order to predict the functional effects of variants across a broad range of ion channels. MissION achieves a significant increase in predictive performance (Area Under the Receiver Operating Characteristic Curve (ROC-AUC): 0.925, compared to 0.897 for the current leading model). Moreover, the model generalizes well to ion channel genes for which little or no electrophysiological recordings are available. MissION provides functional predictions for over 600,000 ion channel variants, made available through an online interface at www.synaptica.nl/variant-interpreter that allows variant interpretation for a wide range of channelopathies.

## 1. Introduction

The human genome contains over 200 genes coding for ion channels that in turn constitute targets for around 18% of FDA-approved drugs [1]. Variants causing dysfunction of the channels encoded by these genes have been associated with at least 130 diseases (channelopathies) affecting various functional systems in the human body, including the nervous [2], cardiovascular [3], and muscular [4] systems, among others. Next-generation sequencing data have allowed a significant rise in the number of identified pathogenic ion channel variants [5]. However, functional classification of these variants comes primarily from labor-intensive patch-clamp experiments, requiring the generation of cell lines expressing the mutated ion channel. Automated patch-clamp setups have improved experimental throughput but still face challenges with respect to seal formation, recording stability, and associated costs [6]. These techniques are cumbersome to apply in a practical setting. As such, new variants are often classified as variants of unknown significance (VUS) and remain a source of uncertainty in the clinical management of the disease [7].

One approach is to preemptively classify thousands of possible variants using so-called multiplex assays of variant effect (MAVE) [8,9], including deep mutational scanning experiments [10]. This approach is expensive and thus far has been applied to only a few ion channel genes [11–13]. Despite the availability of these techniques, reliable variant classification remains one of the biggest short-term challenges to precision genomic medicine [14].

Currently, pathogenicity classification of new variants relies heavily on *in silico* variant effect predictors (VEPs) [15,16], and significant improvements were achieved by using protein language models (pLMs) [17]. However, these predictions do not capture functional changes such as ion selectivity, conductance, and voltage sensing that lead to a gain-of-function (GOF) or a loss-of-function (LOF). G/LOF changes in some ion channels are strongly associated with specific clinical features, e.g., SCN5A (HGNC:10593) GOF variants are often associated with long QT syndrome, whereas SCN5A LOF variants are generally associated with Brugada syndrome [18]. However, in other ion channels, the clinical phenotype does not reliably indicate the underlying functional effect [19,20]. Additionally, some clinically relevant variants may not manifest themselves until later in life [21–23]. Knowing the functional effects of a specific genetic variant can enhance diagnostic accuracy, inform appropriate therapeutic selection, and prevent potential adverse drug reactions due to contraindications [24,25].

A number of machine learning models leveraging existing knowledge on ion channel genetics and electrophysiology have recently been proposed for predicting functional effects of ion channel variants. Models utilizing biophysical [25,26], structural [27], and clinical features [28] have shown promise in accurately predicting the functional effect of variants in ion channels, and have been applied to the whole genome [29,30]. Despite these advances, the overall performance of existing functional effect prediction models lags behind the state-of-the-art VEPs. Additionally, existing approaches often rely on a specific set of manually selected features, so their continued use depends on keeping these tools accessible and up to date online.

To overcome these limitations, we have tested the utility of latent representations from pLMs for the functional effect prediction of channelopathy variants. Similar to large language models trained on extensive language corpora [31], pLMs are trained on large protein sequence datasets and learn latent information embedded in amino acid sequences [32]. We find that pLM representations alone yield strong predictive performance (Area Under the Receiver Operating Characteristic Curve (ROC-AUC): 0.83, Acc: 0.78), and that incorporating gene ontology (GO) terms provides predictive power beyond any existing mode of action models. Our simplified approach matches and surpasses the state-of-the-art models [27,28] that relied on extensive feature sets aggregated from various prediction tools. Notably, the ability of our model to classify variants from out-of-sample genes makes it a promising tool for interpreting variants observed in less well-studied ion channel genes. By providing accurate functional predictions for ion channel variants, our approach can improve diagnostic accuracy, clinical decision-making, and enable more personalized therapeutic strategies.

## 2. Materials and Methods

### 2.1. Dataset assembly

Our dataset comprises 3176 missense variants across 47 ion channel genes, each functionally characterized as either LOF or GOF based on patch-clamp experiments. Each entry includes the gene name, original and substituted amino acid, the residue position of the substitution, the functional effect class (G/LOF), and metadata detailing the source article that provided the functional characterization. This dataset was primarily constructed by integrating data from prior studies [25–28] that employed similar classification algorithms to predict G/LOF outcomes of missense variants. We expanded upon these existing datasets by incorporating recent patch-clamp studies of additional channel genes. To enhance data quality, we implemented a three-step refinement process: (1) removal of duplicate variants, (2) resolution of conflicting classifications using the Functional Epilepsy Nomenclature for Ion Channels (FENICS) [33] as a reference standard, and (3) exclusion of variants where classification conflicts could not be resolved manually.

### 2.2. Variant annotations

We annotated the missense variants following the protocol described in [28], except for eight features where the original publication lacked replicable sources. We augmented this initial set of features using additional sources of annotations, and our complete feature set (Supplementary Table 1) consists of 68 distinct features from various sources, including structural features, topological properties (protein segment and domain information from UniProt), amino acid physicochemical properties, conservation scores, and accessibility features. Beyond these Sequence-, Structure and Residue-based features (hereafter referred to as SSR features), we extracted variant-specific Human Phenotype Ontology (HPO) terms from ClinVar variants with associated Online Mendelian Inheritance in Man (OMIM) identifiers. We mapped OMIM terms to HPO terms using the reference table from NCBI’s MedGen portal [34]. For the variants with clinical annotations, this mapping yielded an average of 18.4 HPO terms per variant, encompassing 783 unique terms across all variants. We further expanded each variant’s HPO term set by including parent terms, resulting in an average of 60.4 terms per variant and 1638 unique terms across the dataset.

For our dataset of variants, we extracted embeddings from the Evolutionary Scale Modeling (ESM) family of pLMs (8M, 12M, 35M, 150M, 650M, 3B), excluding the 15B model due to size and compute resource constraints. The original study reports that performance on various downstream evaluation metrics does not increase significantly between the 3B and 15B models [35]. Therefore, except when indicated otherwise, we used the 3B model in our experiments. We used the canonical sequence of each protein (obtained from UniProt) and applied the substitution as described by the specific missense variants. We then ran each modified sequence through the inference mode of the ESM-2 model and stored the final 2D output representation, which we will refer to as pLM embeddings. Unlike prior versions of the ESM models, ESM-2 does not limit input sequence length, allowing the extraction of full-length sequence embeddings.

### 2.3. Model architecture

Our model processes the pLM embeddings and SSR/HPO/GO features through two separate channels before combining them for final classification (model overview shown in Figure 1). The HPO and GO features are first encoded into multi-hot representations, and then concatenated together with the numerical SSR features. The combined data is then normalised using a z-score standard scaler, and processed through a Multi-Layer Perceptron (MLP - with two hidden layers, each using LeakyReLU activation and dropout).

**Figure 1.**
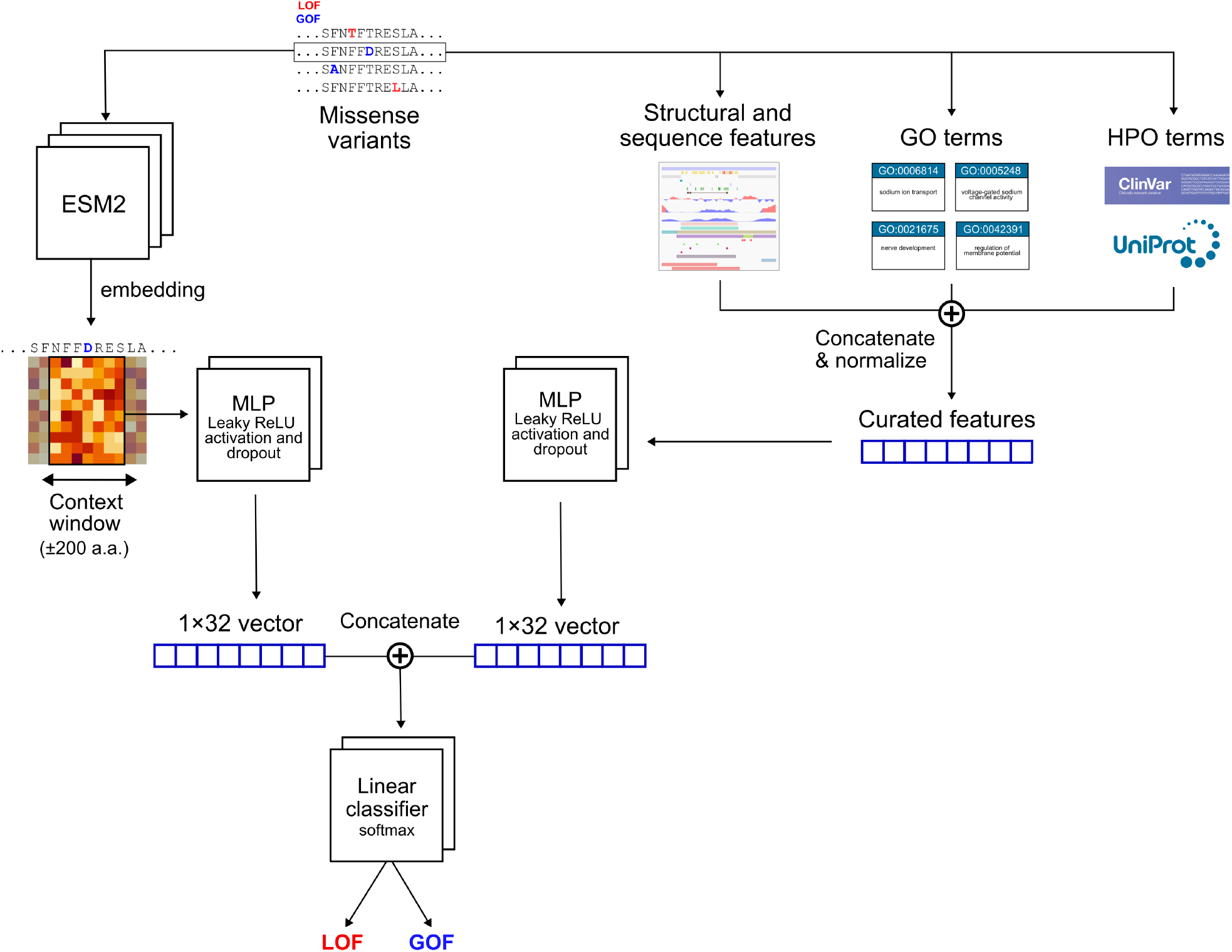
MissION architecture and data flow. MissION integrates diverse data sources and a neural network model to predict the functional effects (G/LOF) of missense variants in ion channel genes. The workflow begins with a curated dataset of missense variants with experimentally determined functional effects. These variants are annotated with structural and sequence-based features, phenotype annotations (mapped to HPO terms), semantic protein representations derived from pLMs, and GO terms. The neural network processes the pLM embeddings and annotated features independently before combining them for a final classification step.

The pLM embeddings have a shape of (N,M), where N denotes the number of residues in the protein sequence, and M represents the embedding dimension, which varies depending on the specific ESM model. To focus on the region surrounding a substitution, the pLM embeddings were cropped to a window of 200 residues on either side of the substitution site, resulting in a matrix of shape (C,M), where C=401 accounts for the window size and includes zero-padding if the substitution is near the start or end of the sequence. This window size can be reduced to ±10 residues without a significant reduction in performance (Supplementary Figure 1). The cropped embedding matrix was then processed through an MLP consisting of three fully connected layers, each followed by a LeakyReLU activation and a dropout layer. The MLP operated on each residue embedding, reducing the dimensionality of each column from M to 16, yielding a matrix of shape (C,16). This matrix was flattened into a vector of length C × 32 and passed through the final MLP layer to produce a latent representation of length 32. This latent vector was concatenated with the latent output vector of the feature’s MLP and fed into a final linear layer with a softmax activation to produce the model’s output.

The model was trained to minimise the focal-loss objective [36]. As opposed to simple class-weighted

cross-entropy, the focal-loss criterion places additional emphasis on hard-to-predict samples by scaling down the loss of easily classified samples. By down-weighting easily classified samples with focusing parameters *γ*=2 and *α*=0.25, focal loss prioritized learning from rare variants and underrepresented classes, offering a theoretical advantage over standard or class-weighted cross-entropy.

Our model was trained with the Adam optimizer regularized through standard weight decay (*α*=10^-4^). During k-fold cross-validation, we trained the model for 20 epochs, using a learning rate of 10^-4^ and a cosine annealing scheduler which reduces the learning rate to 10^-5^ for the final epoch. For each fold, we trained the model across multiple epochs and identified the epoch with the lowest validation loss. We computed performance metrics using the model parameters from this optimal epoch, then averaged these metrics across all folds.

### 2.4. Model evaluation

Evaluation of our model was done primarily through cross-validation – either Repeated Stratified K-Fold or Grouped K-Fold. For Stratified K-Fold, we used a 10 split approach with three repetitions. For Grouped K-Fold, variants from each gene were split into individual folds to evaluate generalisation to the held-out variants, resulting in (k=47) splits. To evaluate performance, we computed several standard metrics, including the ROC-AUC, the Area Under the Precision-Recall Curve (PR-AUC), Matthews Correlation Coefficient (MCC), accuracy, and the F1-score. For the F1-score, we computed the weighted, macro, and binary scores, which place emphasis on different quadrants of the confusion matrix. We compared our model performance against the prior work by Boßelmann et al. [27,28]. For this, we translated their R-lang implementation to Python using comparable implementations (scikit-learn, version 1.4.2) of the used methods. We then re-trained their approach on exactly the same data splits as used to train our Missense ION (MissION) channel variant predictor, and computed the same metrics.

MissION was implemented using Python 3.9.16 and PyTorch 2.3.0. Training was done using CUDA 12.2 on a GTX 1080 GPU, while the pLM embeddings were extracted by running the model using torch hub on an RTX 4090. For evaluation, we used methods from the Python scikit-learn library.

## 3. Results

### 3.1. Ion channel variant dataset and feature distribution

As a first step towards building our machine learning model, we compiled a functional missense variant dataset from the literature. This dataset predominantly features variants from three major ion channel families: sodium, potassium, and calcium (Figure 2A). The balance of LOF and GOF variants varied across different genes, with some genes having variants belonging to only a single functional effect class (Figure 2B). To examine the information carried by the biophysical, structural, sequence, and phenotypic features, we determined their corresponding distribution across the LOF and GOF outcome classes. At the amino acid substitution level, no single substitution was significantly enriched in either class, except for the isoleucine (I) to valine (V) substitution, which showed a significant association with GOF variants (Figure 2C). Next, we investigated the differential distribution of LOF and GOF variants in relation to conserved structural domains across the ion channel family. We aligned the amino acid sequences of all ion channels included in this study to establish a common position system for variant mapping (Figure 2D) using a multiple sequence alignment (MSA) algorithm [37]. As anticipated, the majority of these pathogenic variants were located within the conserved homologous domains (D1-4). Overall, the distribution of variants across these domains did not significantly differ between the LOF and GOF classes (Kolmogorov-Smirnov test: KS statistic=0.076, p=0.71). However, specific features, such as localization within the pore, cytoplasmic, or extracellular domains, exhibited a strong association with the functional effect classes (Figure 2E). Next, we analyzed the information content of HPO terms, which are known to provide substantial information regarding the functional effects of ion channel variants [28]. Our analysis revealed a clear segregation of HPO terms along the LOF-GOF axis (Figure 2F). Furthermore, the majority of HPO terms were associated with only a subset of the genes in our dataset (Figure 2G). This observation confirms that HPO terms serve as a rich source of information about the functional effect of variants, and also carry significant information related to gene identity.

**Figure 2.**
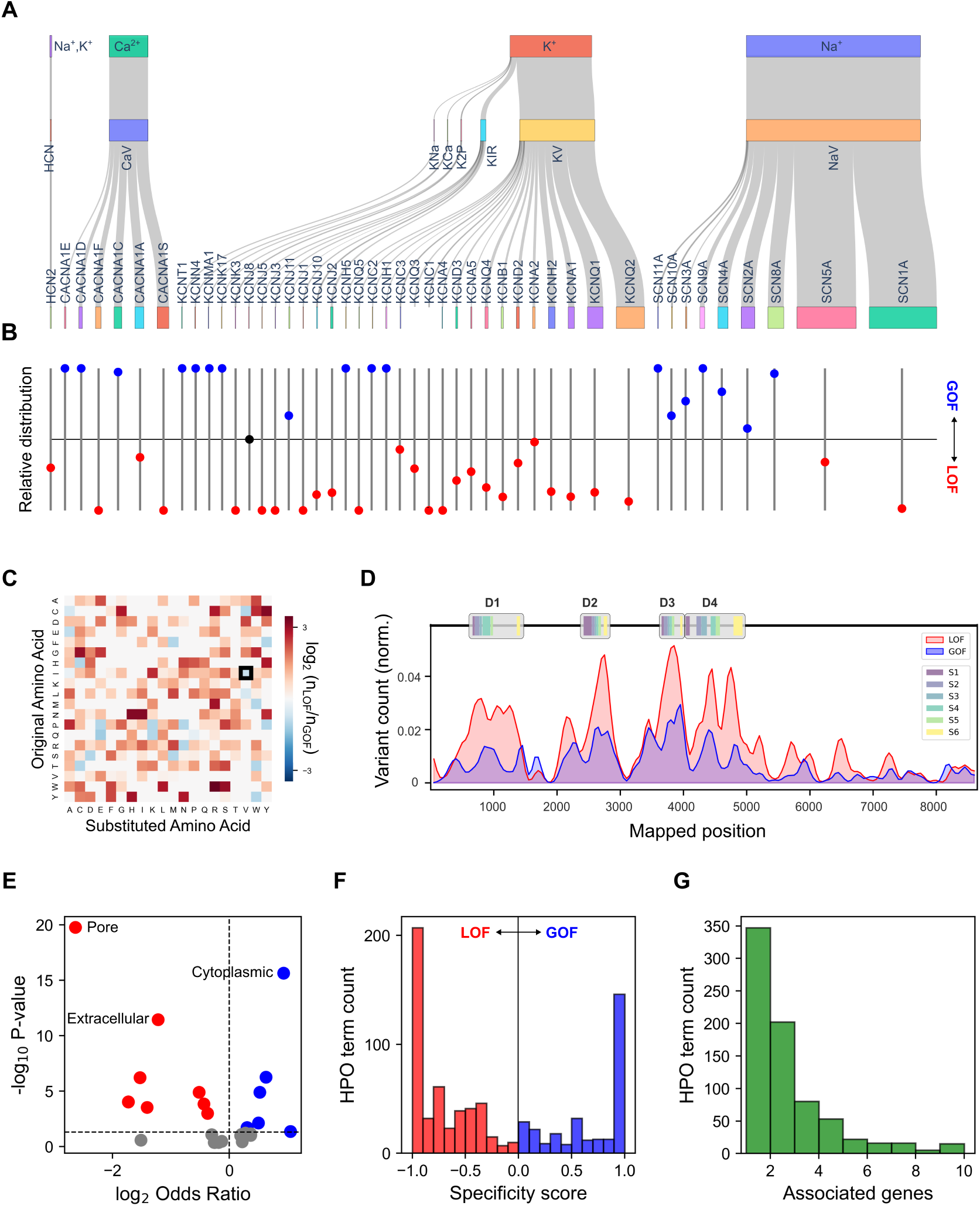
Ion channel variant dataset. **A** Distribution of our dataset of 3176 variants across 47 genes and 8 ion channel families. Each gene is represented by a colored ribbon, whose width indicates the relative number of variants from that gene. Together, the genes are clustered into their respective ion channel families. **B** The ratio of GOF and LOF variants per gene. **C** Only isoleucine (I) to valine (V) showed significant enrichment in GOF variants compared to LOF variants (log_2_(n_LOF_/n_GOF_) ratio shown as heatmap; chi square test for I>V transition, FDR-corrected, p = 0.015). **D** The distribution of variants across subdomains obtained from Multiple Sequence Alignment (MSA) is not significantly different between LOF and GOF classes. The majority of variants are located within regions corresponding to the four homologous domains (D1-4), which contain six transmembrane regions (S1-6). **E** Structural features such as mutated region (pore, extracellular vs cytoplasmic) are significantly enriched in GOF (blue dots) or LOF (red) variants. **F** HPO terms correlate with LOF and GOF effects in ion channel variants (n = 1527 variants with HPO terms). The histogram displays the distribution of HPO term specificity, which is quantified as the difference between GOF and LOF variant counts divided by the total variant count for each term. **G** The majority of HPO terms in our dataset were associated with a limited number of genes.

### 3.2. MissION outperforms the current state-of-the-art model

MissION demonstrates strong predictive performance among the diverse set of ion channels included in our dataset. To evaluate model performance given a widespread class imbalance in our dataset, we used the ROC-AUC and PR-AUC as the main performance metrics (see Methods for details). ROC-AUC provides a robust measure of the model’s ability to discriminate between LOF and GOF variants, whereas PR-AUC captures the model’s ability to predict GOF variants that are less common in the dataset. Cross-validated performance of MissION reached an ROC-AUC of 0.925, with a PR-AUC of 0.865 (Table 1). This performance surpasses that of the SCION model, a prior multi-task learning (MTL) framework [27], which employed a comparable set of curated features in combination with a support vector machine for classification (Figure 3A).

**Table 1.** Comparison of MissION and SCION. Repeated stratified k-fold cross-validation (k=10, n=3) comparing our MissION model against a previous state-of-the-art approach. Values shown in bold represent statistically significant increases as determined by paired t-test.

| Dataset | Model | ROC-AUC | PR-AUC | MCC | F1-weighted | F1-macro | F1-binary | Accuracy |
| --- | --- | --- | --- | --- | --- | --- | --- | --- |
| Full<br>($n=3176$) | MissION | **0.925**<br>$\pm 0.017$ | **0.865**<br>$\pm 0.027$ | **0.653**<br>$\pm 0.044$ | **0.854**<br>$\pm 0.018$ | **0.824**<br>$\pm 0.023$ | 0.749<br>$\pm 0.034$ | **0.857**<br>$\pm 0.018$ |
| | SCION | 0.897<br>$\pm 0.021$ | 0.815<br>$\pm 0.038$ | 0.612<br>$\pm 0.046$ | 0.827<br>$\pm 0.021$ | 0.802<br>$\pm 0.023$ | 0.737<br>$\pm 0.030$ | 0.838<br>$\pm 0.022$ |
| Phenotype-<br>annotated<br>($n=1528$) | MissION | 0.970<br>$\pm 0.011$ | 0.935<br>$\pm 0.026$ | 0.820<br>$\pm 0.051$ | 0.928<br>$\pm 0.021$ | 0.909<br>$\pm 0.026$ | 0.866<br>$\pm 0.029$ | 0.929<br>$\pm 0.020$ |
| | SCION | 0.964<br>$\pm 0.014$ | 0.929<br>$\pm 0.032$ | 0.812<br>$\pm 0.037$ | 0.923<br>$\pm 0.016$ | 0.905<br>$\pm 0.019$ | 0.864<br>$\pm 0.027$ | 0.923<br>$\pm 0.016$ |

**Figure 3.**
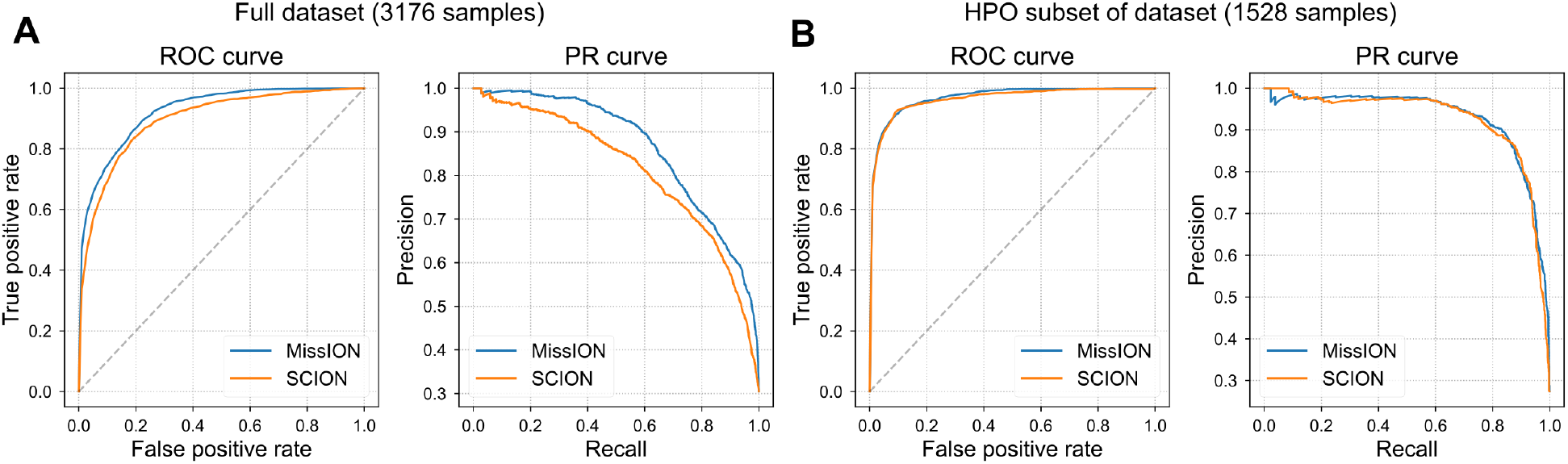
MissION outperforms existing SCION model on the full dataset. **A** Performance comparison on the complete dataset without HPO features (MissION ROC-AUC: 0.925 ± 0.017, significantly higher than SCION ROC-AUC: 0.897 ± 0.021), and **B** with the inclusion of HPO terms as input features and only using the subset of the data for which HPO terms are available (MissION ROC-AUC: 0.970 ± 0.011; SCION ROC-AUC: 0.964 ± 0.014).

Next, we incorporated HPO terms to benchmark MissION against another version of the SCION model, utilizing a multi-kernel framework (MKL) [28]. Although HPO terms were only available for about half of the variants in our dataset, inclusion of these annotations in classification led to significantly better performance for both models, with MissION reaching an ROC-AUC score of 0.970 and PR-AUC of 0.935 (Figure 3B). This result is consistent with our exploratory observations (Figure 2F-G) and previous findings [28,38], which demonstrated that clinical phenotypes are highly informative about the functional outcomes of observed variants.

Model accuracy and prediction confidence are crucial for clinical and research applications. To evaluate this aspect of our model, we examined the relationship between its raw probability outputs and the true labels. We treat the model output probabilities for each class label as a proxy confidence value in its prediction. We then assessed the calibration of these probabilities by constructing a calibration curve. We binned the model’s GOF scores from 10-fold cross-validation into 10 equal intervals and computed the frequency of true GOF variants within each bin (Figure 4A). To characterize the calibration of our model, we computed the precision for a range of MissION score thresholds (as described in [28]) and determined the thresholds that achieve 90% precision per class (Figure 4B). To further quantify the prediction quality, we examined how error rates varied as a function of model confidence for both LOF and GOF predictions by binning their respective probabilities (Figure 4C). We observe that error rates declined as prediction confidence increased, indicating good predictive capability. Based on these error rates and the clustering of predictions around 0 and 1, we suggest using more conservative classification thresholds: 0.1 for LOF and 0.9 for GOF, and to label the variants in between as uncertain.

**Figure 4.**
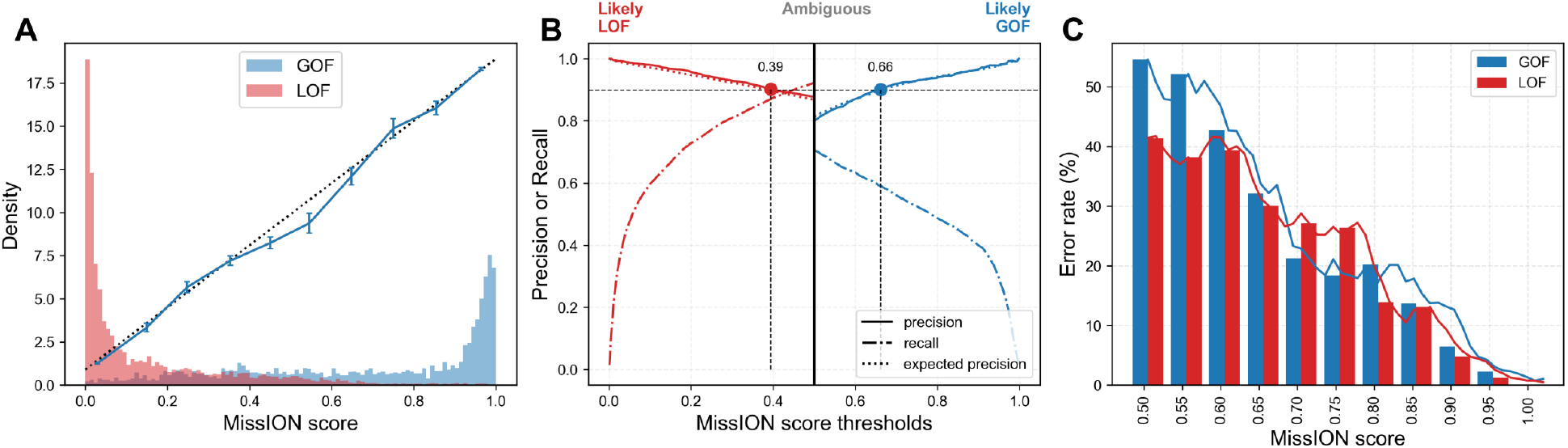
Prediction confidence and calibration analysis. **A** MissION score, here representing the probability of GOF classification and colored according to the true label (GOF or LOF). The blue calibration curve shows the model’s reliability by relating the average predicted GOF probability to the observed fraction of GOF cases. A perfectly calibrated model would align with the dotted diagonal line. **B** Precision and recall for MissION predictions across cross-validation folds, computed for each output class. GOF samples are plotted in the [0.5,1] range, while LOF samples are plotted in the [0, 0.5] range, for which the MissION score is set as 1 - LOF probability. The red and blue dots indicate the 90% precision threshold. The dotted line (expected precision) shows the predicted probability of a variant being correctly labeled as LOF or GOF, respectively. **C** Error rate of predictions as a function of the MissION score. Bars represent the error rate within specific score intervals, while the lines show a moving average of the error rate (window size = 0.05).

### 3.3. MissION shows the highest generalization performance on out-of-sample genes

Classification models trained on class-imbalanced datasets are susceptible to the ‘Type 2’ data circularity problem [39], where models learn gene-specific class distributions rather than functional patterns. This occurs when labeled variants within individual genes exhibit extreme class homogeneity - for example, when nearly all characterized variants in a gene are either LOF or GOF. Under these conditions, models may simply learn gene identity rather than develop generalizable classification rules. This limitation is particularly problematic in cases where many genes contain few variants. To assess MissION’s ability to make accurate predictions on unrepresented or underrepresented genes, we performed a Grouped K-Fold cross-validation experiment, in which all variants from a single gene were held out as the validation set in each fold (zero-shot learning).

Overall accuracy of MissION was 70.8% (Figure 5), while SCION performed at 62.6% accuracy (two-sided paired t-test *t*(46)=1.04 *p*=0.17). Both models significantly outperformed chance-level (two-sided paired t-tests: *t*(46)=4.02, *p*=2.1×10^-4,^ and *t*(46)=2.43, *p*=0.019, respectively). Despite the expected drop in overall performance, for 39 out of 47 genes, the accuracy was above chance level. Accuracy for out-of-sample genes varied widely across genes and models, suggesting the need for an ensemble approach for handling cases where genes have few known variants.

**Figure 5.**
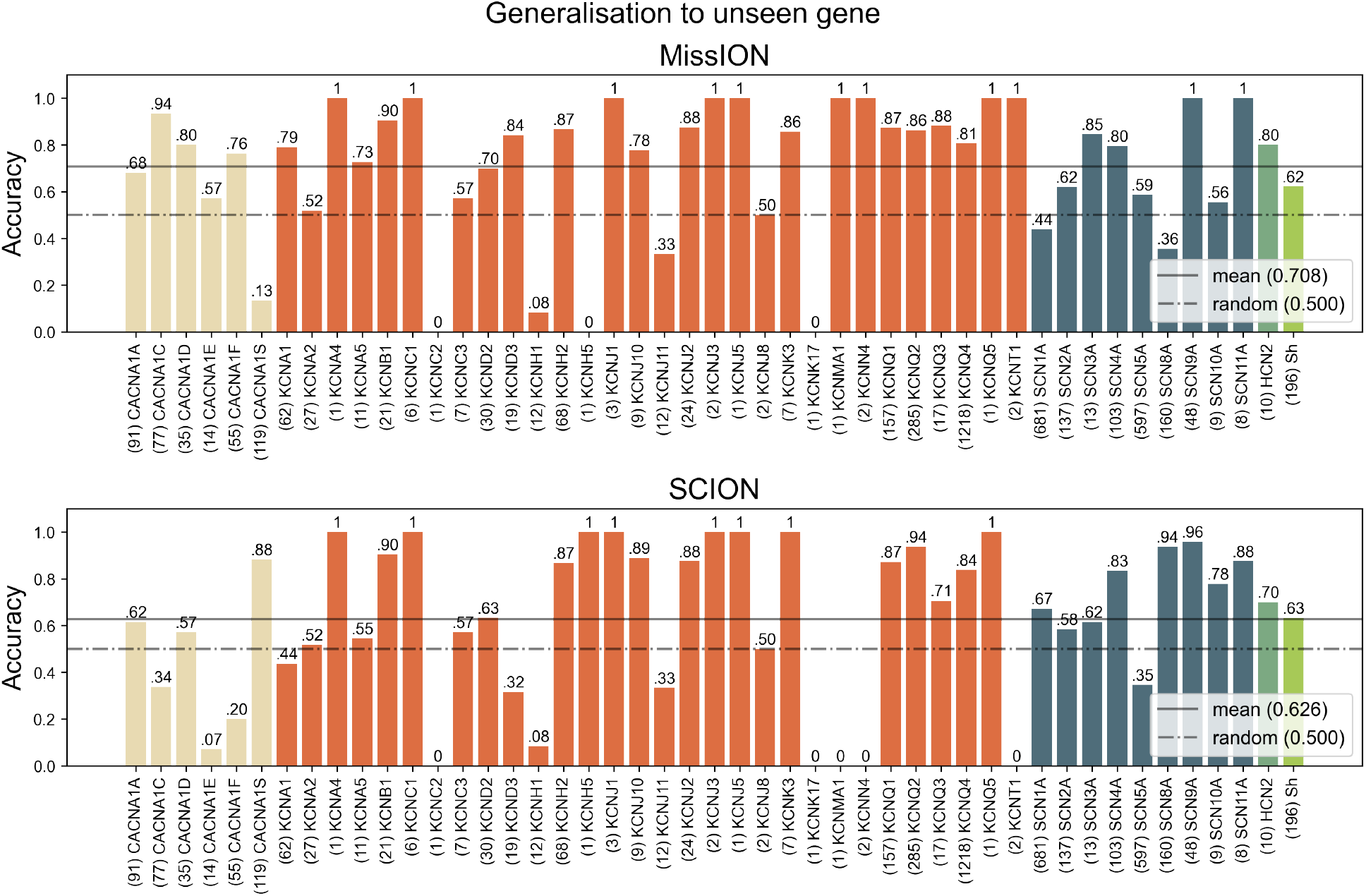
Cross-gene generalization via zero-shot learning. The model’s performance was assessed on variants of genes entirely excluded from the training data (zero-shot setting). Bar heights show accuracy for each tested gene, with colors indicating their respective ion channel families. The number of variants within each gene is shown in parentheses. Performance of a uniform random classifier is shown as a dotted line for comparison, while the average MissION and SCION performances are shown as a solid line.

### 3.4. The full MissION model outperforms ablated and isolated versions of the model

To understand the contribution of each information source to MissION’s predictive performance, we conducted ablation and isolation experiments and quantified the performance of the resulting reduced models (Supplementary Table 2). In the ablation experiments, we systematically withheld one of the three annotation sources (pLM, GO, and SSR sets) during model training and evaluation. This methodology enabled a comparative analysis of the individual information sources and an assessment of their potential synergistic effects. Complementarily, the isolation experiments involved training and evaluating models using only a single specified information source.

The ablation experiments revealed a significant performance decrease for all evaluation metrics upon the exclusion of GO terms (*t*(29)=10.68 *p*=1.5×10^-11^). In agreement with a previous study [27], this observation strongly suggests that the model benefits from learning gene-specific variant distributions and inter-gene similarities relevant to the task. This finding is further supported by the highest performance of the GO only model compared to any other isolated model (one-way ANOVA: F = 250.5, p = 7.9 × 10*−*^37^; post-hoc Tukey HSD tests: GO vs. pLM, mean difference = 0.055, p = 1.6 × 10*−*^14^; GO vs. SSR, mean difference = 0.128, p =2.2×10^*−*15^; pLM vs. SSR, mean difference = 0.074, p =2.2×10^*−*15^).

Statistical comparison of ablated models against the full model using Dunnett’s test revealed that only removing GO features resulted in significantly reduced performance (GO removal: mean difference = -8.827, p=9.2 × 10*−*^15^; pLM removal: mean difference = -1.016, p=0.61; SSR removal: mean difference = -2.218, p=0.074). These results indicate that GO features provide unique, essential information that cannot be compensated by other feature types. In contrast, pLM and SSR features appear to be largely redundant with each other, suggesting these feature types capture overlapping information rather than providing distinct predictive signals.

## 4. Discussion

The application of evidence-based practice currently relies largely on the observed clinical phenotype. For channelopathies, among other conditions, understanding the underlying genetic causes can aid in treatment selection and help identify potential contraindications. Gene-level information is insufficient; it is crucial to study the consequences of the observed variant. For example, a child with a de novo variant in SCN2A (HGNC:10588) presenting with seizures before 3 months of age is likely to have a GOF variant, whereas onset after 3 months is likely to be caused by a LOF variant, and will respond differently to sodium channel blockers, making treatment decisions difficult [23,40]. Complicating matters further, LOF and GOF variants in the same gene may lead to similar phenotypes, as demonstrated by certain SCN8A (HGNC:10596) variants that cause epileptic encephalopathy [41]. The absence of phenotypic data is also possible. For example, asymptomatic LOF variant carriers in KCNH2 (HGNC:6251) can have a higher risk of cardiac events, warranting the administration of beta blockers once such a variant is found [42]. The need to address cases like these drives the development of novel machine learning models that can readily inform clinical variant interpretation.

Here, we showed state-of-the-art functional outcome predictions for ion channel variants. This was related to two major advances: 1) expansion of the variant dataset to 3176 missense variants across 47 genes from three major ion channel families (K^+^, Na^+^, and Ca^2+^), creating the largest collection of functionally characterized ion channel variants to date, and 2) integrating two novel sources of information, namely: protein evolutionary conservation through pLM embeddings and gene-level semantic knowledge through GO terms. Currently, accurate classification of ion channel variants remains difficult due to two main challenges: severe G/LOF class imbalances within genes and limited training data available for a majority of ion channel genes. This class imbalance may partly reflect inherent biophysical constraints, as different structural domains show differential susceptibility to GOF vs LOF variants, though sampling and study biases also likely contribute [39]. We addressed the imbalance through the focal-loss objective by down-weighting easy examples and emphasising hard-to-classify cases, thereby prioritizing learning from underrepresented classes. To deal with differences in data availability per gene, we performed zero-shot learning in order to assess performance more evenly for all genes. Variability in performance between the models tested also suggests that it may be beneficial to use an ensemble of models, taking into account prediction confidences, to optimize performance across genes.

Beyond the modelling challenges discussed above, the quality of ground truth data presents additional complications. Unlike the pathogenicity classification task, where established certifications and guidelines are established to weigh different lines of evidence, ground truth data for ion channel functional effect models come directly from patch-clamp experiments. While the outcome of a variant can typically be inferred directly through experimental recordings, classification is sometimes complicated by factors such as the expression system used in the experiment [43], or mixed effects that are seen in some variants [44]. Furthermore, not all relevant biophysical parameters are reported in many publications. We addressed some of these complications by applying the newly proposed FENICS ontology [33] for variants with conflicting reported functional outcomes, and by excluding variants where resolution was not possible. Future studies utilizing high-throughput automated patch-clamp recordings and standardized experimental protocols, along with systematic reporting of measured parameters, will likely improve data quality and classification performance [23].

Having established the value of combining multiple information modalities in our model, we then analyzed the specific contribution of each modality in order to enhance the interpretability of the model, which is critical in clinical and academic settings [45]. Previous modelling approaches in this field have primarily relied on a long list of specially curated features derived from various sources and tools [27,28]. While the best-performing model incorporates all modalities, excluding the list of curated (SSR) features showed comparable performance, providing opportunities for simplified applications.

The success of our multimodal framework builds upon the evolutionary characteristic that the majority of ion channel types found in humans first appeared in prokaryotes and exhibit common core structural motifs throughout evolution [46], making them particularly amenable to computational analysis. As such, many studies focus on these conducting parts of the ion channels, often excluding accessory subunits, which can also contain pathogenic variants [47]. Using newly improved multimeric protein structure prediction capabilities [48], future models utilizing variant-specific protein structures could help address this limitation and may also help extend to classify variant types other than missense variants (e.g., insertion and deletions that make up 15-21% of all variants [49]).

In conclusion, we introduce MissION, a machine learning classifier that predicts the functional effects of missense variants in ion channels by integrating protein language model embeddings, biophysical features, and gene-level knowledge. MissION enables the currently most accurate GOF/LOF classification for over 600,000 ion channel variants through an online portal, available at www.synaptica.nl/variant-interpreter, offering clinically relevant insights that can guide diagnosis and treatment in channelopathies without relying on patch-clamp experiments.

## 5. Declarations

### 5.1. Data Availability

The datasets of functional predictions generated during this study are available through the Synaptica Variant Interpreter (www.synaptica.nl/variant-interpreter). The manuscript is currently under review. Upon publication, the full implementation of the code will be made available on GitHub.

### 5.2. Funding Statement

The research in this manuscript was funded by Synaptica Ltd., with the supporting grant MIT-R&D- Artificiële Intelligentie (AI) 2023 by the Dutch Ministry of Commerce (RVO), grant number MIT-AI-23-03474124.

### 5.3. Author Contributions

Conceptualization: AA, SG, MBM. Data curation: AA. Funding acquisition: MBM. Investigation: AA, SG, MBM. Methodology: SG. Software: AA, SG. Visualization: AA, SG. Writing: AA, SG, MBM, PHET

### 5.4. Conflicts of Interest

This research presented here was performed by Synaptica Ltd. Author MBM is the founder of Synaptica, and authors SG and AA are employees of Synaptica. There are no competing interests for author PHET (Professor at Radboud University Nijmegen, the Netherlands).

## Notes

### Clinical Protocols

https://www.synaptica.nl/variant-interpreter

### Funding Statement

The research in this manuscript was funded by Synaptica Ltd., with the supporting grant MIT-R&D-Artificiele Intelligentie (AI) 2023 by the Dutch Ministry of Commerce (RVO), grant number MIT-AI-23-03474124.

### Author Declarations

This study uses ONLY openly available data that from the scientific and medical literature.

