## Supplementary Material for "Functional Effect Predictions For Ion Channel Missense Variants Using a Protein Language Model"

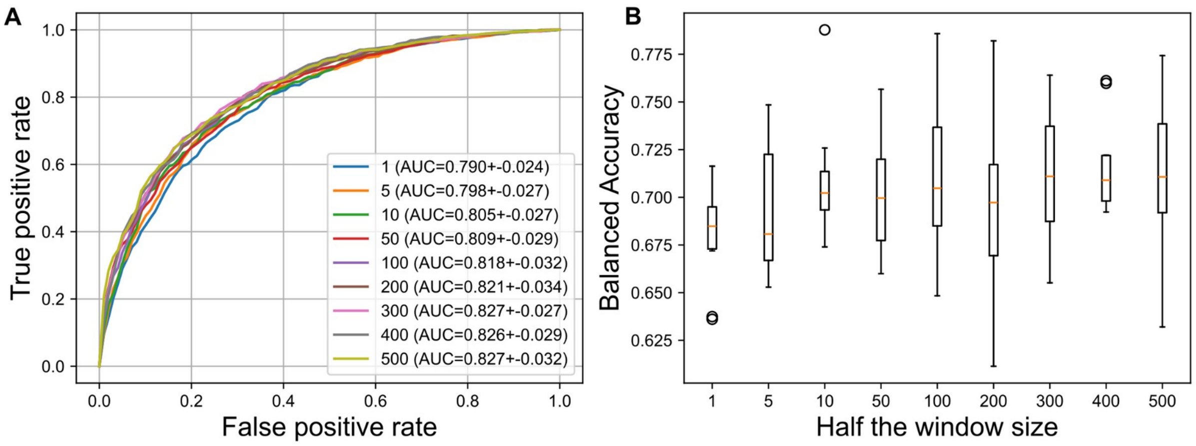


**Supplementary Figure 1: pLM-only model performance across different window sizes.** Increasing the number of residues around the substituted residue that are included in the model has a non-significant effect on performance when only pLM features are used. An Anova and post-hoc Tukey HSD were performed for all measured metrics. The only significant difference found was for the PR-AUC metric for the 1 residue window size (Tukey-HSD(0.0729) *p*=0.0277). **A.** The ROC curves at different half-window sizes, averaged across 10 k-fold (k=10, n=3) cross validations. **B.** The window size does also not significantly impact balanced accuracy.

**Supplementary Table 1. Feature set including structural and topological properties.**

Predictors (n=68) used in training MissION. Values of predictors related to amino acid transitions (BLOMAP, BLOSUM62, etc.) are taken from their respective reference tables.

| PREDICTOR | DESCRIPTION | TYPE | SOURCE |
|---|---|---|---|
| aa_blomap_aa1_dim1 | Encoding of the physicochemical properties of the original amino acid using a Naïve Bayes classifier, first encoding dimension | Numerical | Reference table |
| aa_blomap_aa1_dim2 | Encoding of the physicochemical properties of the original amino acid using a Naïve Bayes classifier, second encoding dimension | Numerical | Reference table |
| aa_blomap_aa1_dim3 | Encoding of the physicochemical properties of the original amino acid using a Naïve Bayes classifier, third encoding dimension | Numerical | Reference table |
| aa_blomap_aa1_dim4 | Encoding of the physicochemical properties of the original amino acid using a Naïve Bayes classifier, fourth encoding dimension | Numerical | Reference table |
| aa_blomap_aa1_dim5 | Encoding of the physicochemical properties of the original amino acid using a Naïve Bayes classifier, fifth encoding dimension | Numerical | Reference table |
| aa_blomap_aa2_dim1 | Encoding of the physicochemical properties of the substituted amino acid using a Naïve Bayes classifier, first encoding dimension | Numerical | Reference table |
| aa_blomap_aa2_dim2 | Encoding of the physicochemical properties of the substituted amino acid using a Naïve Bayes classifier, second encoding dimension | Numerical | Reference table |
| aa_blomap_aa2_dim3 | Encoding of the physicochemical properties of the substituted amino acid using a Naïve Bayes classifier, third encoding dimension | Numerical | Reference table |
| aa_blomap_aa2_dim4 | Encoding of the physicochemical properties of the substituted amino acid using a Naïve Bayes classifier, fourth encoding dimension | Numerical | Reference table |
| aa_blomap_aa2_dim5 | Encoding of the physicochemical properties of the substituted amino acid using a Naïve Bayes classifier, fifth encoding dimension | Numerical | Reference table |
| aa_blosum62 | amino acid substitution value according to the midrange Blocks Substitution Matrix (BLOSUM62) | Numerical | Reference table |

| | | | |
|---|---|---|---|
| aa_braun_aa1_E1 | Encoding of the physicochemical properties of the original amino acid using multidimensional scaling, first encoding dimension | Numerical | Reference table |
| aa_braun_aa1_E2 | Encoding of the physicochemical properties of the original amino acid using multidimensional scaling, second encoding dimension | Numerical | Reference table |
| aa_braun_aa1_E3 | Encoding of the physicochemical properties of the original amino acid using multidimensional scaling, third encoding dimension | Numerical | Reference table |
| aa_braun_aa1_E4 | Encoding of the physicochemical properties of the original amino acid using multidimensional scaling, fourth encoding dimension | Numerical | Reference table |
| aa_braun_aa1_E5 | Encoding of the physicochemical properties of the original amino acid using multidimensional scaling, fifth encoding dimension | Numerical | Reference table |
| aa_braun_aa2_E1 | Encoding of the physicochemical properties of the substituted amino acid using multidimensional scaling, first encoding dimension | Numerical | Reference table |
| aa_braun_aa2_E2 | Encoding of the physicochemical properties of the substituted amino acid using multidimensional scaling, second encoding dimension | Numerical | Reference table |
| aa_braun_aa2_E3 | Encoding of the physicochemical properties of the substituted amino acid using multidimensional scaling, third encoding dimension | Numerical | Reference table |
| aa_braun_aa2_E4 | Encoding of the physicochemical properties of the substituted amino acid using multidimensional scaling, fourth encoding dimension | Numerical | Reference table |
| aa_braun_aa2_E5 | Encoding of the physicochemical properties of the substituted amino acid using multidimensional scaling, fifth encoding dimension | Numerical | Reference table |
| aa_grantham | Grantham distance between the original and substituted amino acids, based on composition, polarity and residue volume | Numerical | Reference table |
| aa_hphob_pca1 | change in hydrophobicity by amino acid substitution using 98 hydrophobicity scales and principal component analysis, first component | Numerical | Reference table |
| aa_hphob_pca2 | change in hydrophobicity by amino acid substitution using 98 hydrophobicity scales and principal component analysis, first component | Numerical | Reference table |
| aa_hphob_pca3 | change in hydrophobicity by amino acid substitution using 98 hydrophobicity scales and principal component analysis, first component | Numerical | Reference table |
| B_ss8 | 8-class secondary structure classification (SS8) by NetSurfP – 2.0 and DSSP | Categorical | NETSURF & DSSP |
| C_ss3 | 3-class secondary structure classification (SS3) by NetSurfP – 2.0 and DSSP | Categorical | NETSURF & DSSP |
| C_ss8 | 8-class secondary structure classification (SS8) by NetSurfP – 2.0 and DSSP | Categorical | NETSURF & DSSP |
| str_top_cytoplasmic | position on protein topology via UniProt, cytoplasmic | Categorical | UniProt |
| E_ss3 | 3-class secondary structure classification (SS3) by NetSurfP – 2.0 and DSSP | Categorical | NETSURF & DSSP |
| E_ss8 | 8-class secondary structure classification (SS8) by NetSurfP – 2.0 and DSSP | Categorical | NETSURF & DSSP |
| str_top_extracellular | position on protein topology via UniProt, extracellular | Categorical | UniProt |
| G_ss8 | 8-class secondary structure classification (SS8) by NetSurfP – 2.0 and DSSP | Categorical | NETSURF & DSSP |
| H_ss3 | 3-class secondary structure classification (SS3) by NetSurfP – 2.0 and DSSP | Categorical | NETSURF & DSSP |
| H_ss8 | 8-class secondary structure classification (SS8) by NetSurfP – 2.0 and DSSP | Categorical | NETSURF & DSSP |
| I_ss8 | 8-class secondary structure classification (SS8) by NetSurfP – 2.0 and DSSP | Categorical | NETSURF & DSSP |
| str_top_porehelix | position on protein topology via UniProt, pore | Categorical | UniProt |

| | | | |
|---|---|---|---|
| S_ss8 | 8-class secondary structure classification (SS8) by NetSurfP – 2.0 and DSSP | Categorical | NETSURF & DSSP |
| str_top_S0 | position on protein topology via UniProt, segment S0 | Categorical | UniProt |
| str_top_S1 | position on protein topology via UniProt, segment S1 | Categorical | UniProt |
| str_top_S2 | position on protein topology via UniProt, segment S2 | Categorical | UniProt |
| str_top_S3 | position on protein topology via UniProt, segment S3 | Categorical | UniProt |
| str_top_S4 | position on protein topology via UniProt, segment S4 | Categorical | UniProt |
| str_top_S5 | position on protein topology via UniProt, segment S5 | Categorical | UniProt |
| str_top_S6 | position on protein topology via UniProt, segment S6 | Categorical | UniProt |
| str_asa | change in residue accessible surface area predicted by PROF via PredictProtein | Numerical | PredictProtein |
| str_consurf | probabilistic positional evolutionary conservation estimates by ConSurf via PredictProtein | Numerical | PredictProtein |
| str_helix | prediction of topology for helical transmembrane proteins by PHDhtm_top via PredictProtein, network output for helix | Numerical | PredictProtein |
| str_isis | prediction of residue involvement in protein-protein interaction sites by ISIS | Numerical | PredictProtein |
| str_loop | prediction of topology for helical transmembrane proteins by PHDhtm_top via PredictProtein, network output for loop | Numerical | PredictProtein |
| str_nors | prediction of protein disorder by NORSnet via PredictProtein | Numerical | PredictProtein |
| str_pbie_b | state of residue relative accessible surface area predicted by PROF via PredictProtein, buried | Categorical | PredictProtein |
| str_pbie_e | state of residue relative accessible surface area predicted by PROF via PredictProtein, exposed | Categorical | PredictProtein |
| str_pbie_i | state of residue relative accessible surface area predicted by PROF via PredictProtein, intermediate | Categorical | PredictProtein |
| str_prhl_H | maximum score model prediction of topology for helical transmembrane proteins by PHDhtm_top via PredictProtein, helical membrane | Categorical | PredictProtein |
| str_prhl_L | maximum score model prediction of topology for helical transmembrane proteins by PHDhtm_top via PredictProtein, no helical transmembrane | Categorical | PredictProtein |
| str_pito_i | predicted topology of transmembrane regions by PHDhtm_top via PredictProtein, loop inside | Categorical | PredictProtein |
| str_pito_o | predicted topology of transmembrane regions by PHDhtm_top via PredictProtein, loop outside | Categorical | PredictProtein |
| str_pito_T | predicted topology of transmembrane regions by PHDhtm_top via PredictProtein, transmembrane | Categorical | PredictProtein |
| str_profbval | prediction of protein disorder by PROFbval via PredictProtein | Numerical | PredictProtein |
| str_rsa | change in residue relative accessible surface area predicted by PROF via PredictProtein | Numerical | PredictProtein |
| str_ucon | prediction of protein disorder by Ucon via PredictProtein | Numerical | PredictProtein |
| T_ss8 | 8-class secondary structure classification (SS8) by NetSurfP – 2.0 and DSSP | Categorical | NETSURF & DSSP |
| HPO | HPO terms for this variant | Binary | HPO |
| HPOParents | Parent HPO terms for this variant | Binary | HPO |
| GO | GO terms for this gene | Binary | GO |
| AlphaMissense | raw pathogenicity prediction by AlphaMissense | Numerical | AlphaMissense |
| ESM_LLR | ESM log likelihood ratio | Numerical | ESM |

**Supplementary Table 2. Feature importance analysis.**
Here we compare performance when ablating various features from the model. Performance metrics are averaged across k-fold (k=10, n=3) splits. Italic indicates significantly worse performance compared to the full MissION model, tested by independent t-tests with Bonferroni correction.

| Condition | ROC-AUC | PR-AUC | MCC | F1-weighted | F1-macro | F1-binary | Accuracy |
|---|---|---|---|---|---|---|---|
| MissION | **0.925±0.017** | **0.865±0.027** | **0.653±0.044** | **0.854±0.018** | **0.824±0.023** | **0.749±0.034** | **0.857±0.018** |
| MissION ex. SSR | 0.915±0.017 | 0.846±0.029 | 0.624±0.045 | 0.841±0.019 | 0.808±0.024 | 0.725±0.037 | 0.845±0.018 |
| MissION ex. GO | *0.867±0.021* | *0.759±0.042* | *0.541±0.060* | *0.806±0.025* | *0.767±0.033* | *0.666±0.054* | *0.811±0.023* |
| Mission ex. pLM | 0.918±0.019 | 0.858±0.030 | 0.636±0.050 | 0.846±0.021 | 0.813±0.026 | 0.729±0.039 | 0.851±0.020 |
| SSR only | *0.731±0.026* | *0.517±0.044* | *0.213±0.056* | *0.671±0.021* | *0.583±0.029* | *0.355±0.049* | *0.707±0.018* |
| GO only | *0.887±0.020* | *0.789±0.032* | *0.592±0.049* | *0.823±0.020* | *0.781±0.025* | *0.672±0.039* | *0.835±0.018* |
| pLM only | *0.832±0.034* | *0.700±0.057* | *0.461±0.075* | *0.774±0.031* | *0.727±0.039* | *0.606±0.062* | *0.780±0.028* |
